# Limits of Single-Pass Retrieval-Augmented Generation for AI-Powered Cancer Care Navigation: A Comparison of Retrieval Strategies

**DOI:** 10.64898/2026.08.31.26361774

**Authors:** Emrul Hasan, Yuci Zhang, Olivia Cook, Andrew Loe, Matthew Sha, Leah T’ien, Margaret Ng, Chris Rauscher, Srinivas Raman, Jacqueline L. Bender, Raymond T. Ng, Alan Bates, John-Jose Nunez

## Abstract

**Background:** People affected by cancer often face difficulty finding relevant clinical, psychological, and practical support services. AI-powered navigation assistants may improve access to these resources, but their retrieval performance must be reliable.

**Objective:** To develop a single-pass retrieval-augmented generation assistant for cancer-care navigation and compare the retrieval strategies, including their robustness to reworded questions.

**Methods:** We created a database of 853 cancer-support resources reviewed by librarians, clinicians, researchers, and patient partners. We evaluated the system using 100 questions derived from questions submitted by patients. We compared keyword-based, semantic, and hybrid retrieval using Precision@K, Hit@K, and nDCG@K. The best-performing configuration was then tested using semantically equivalent rewordings of the original questions.

**Results:** Keyword-based retrieval performed poorly, achieving a P@1 of 25.0% and Hit@5 of 43.0%. Semantic retrieval improved these results to 58.0% and 86.0%, respectively. The best hybrid configuration achieved a P@1 of 64.0%, Hit@5 of 90.0%, and nDCG@5 of 51.0%. Performance remained relatively stable when the questions were reworded, with a P@1 of 61.0%, Hit@5 of 88.0%, and nDCG@5 of 46.1%.

**Conclusions:** Hybrid retrieval performed best and remained relatively stable when questions were reworded. However, its limited ability to rank a relevant resource first highlights the limitations of single-pass retrieval for patient-facing cancer navigation. Future work will explore metadata filtering and a multi-agent architecture to improve retrieval reliability.

## Introduction

Despite growing supportive care resources, cancer patients and their families face barriers navigating clinical, psychological, and logistical services [2, 3, 8]. These gaps delay access to care [1], whereas patient navigation improves screening participation, reduces diagnostic and treatment delays, and enhances quality of life and satisfaction [3].

Traditional clinician-, lay-, or peer-led navigation can be difficult to scale given workforce shortages, attrition, and budget constraints [7]. AI-powered conversational assistants may address this gap by delivering personalized resource recommendations. To our knowledge, no AI navigation assistant has been designed for cancer patients generally, though related tools exist for specific cancer types [10].

In this study, we developed an AI-powered cancer navigation assistant that uses single-pass retrieval-augmented generation (RAG): for each question, the system retrieves matching resources from a curated database, then generates a recommendation. We conducted a comparative evaluation study of retrieval strategies, then tested whether the best strategy held up when questions were reworded (metamorphic testing) [4].

## Methods

### Resource Database and Indexing

We constructed a database of 853 BC Cancer library-approved or patient partner-recommended resources, each with a human-annotated description. The database was reviewed by clinicians, researchers, and patient partners. Descriptions were embedded using Qwen3-Embedding-0.6B [9], capturing their semantic meaning.

### Development of Question-Answer Pairs

We built a development dataset of 100 question-answer (QA) pairs derived from questions patients submitted at the BC Cancer Summit or to the BC Cancer Library, spanning all major cancer sites (Figure 1). This study was approved by the UBC BC Cancer Research Ethics Board. For confidentiality, research assistants rewrote each question, altering geographic and demographic details while preserving intent. The resulting pairs were reviewed by three patient partners for plausibility and verified by a clinical expert (Multimedia Appendix 1).

**Figure 1.**
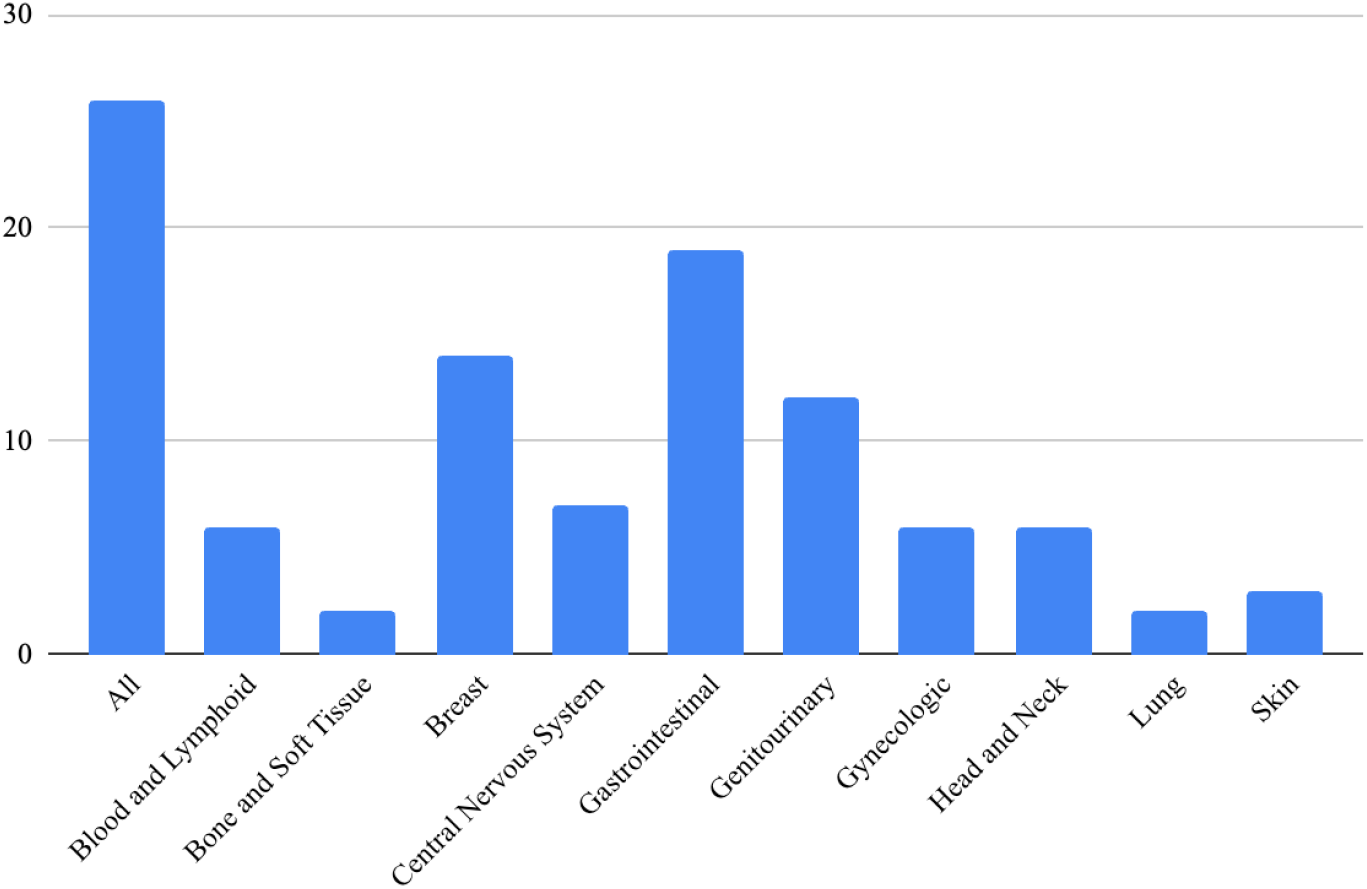
11 distinct question categories by cancer type (n=100). Questions with multiple cancer type tags are counted for each category.

### Resource Retrieval Methods

The system retrieves resources using dense (semantic) retrieval, which encodes each query with the same embedding model and matches resource descriptions by k-nearest neighbour search, or sparse (keyword) retrieval with BM25 [6], which searches the resource title and document text fields, weighting titles two-fold. We compared three retrieval methods: dense, sparse, and hybrid combining both result lists with Reciprocal Rank Fusion (RRF; Multimedia Appendix 2). Retrieved resources are deduplicated and normalized, and the top K are assembled into a prompt for a large language model (Llama 3 70B Instruct) [5] to generate the final response.

### Evaluation

To evaluate our system, we applied metamorphic testing, in which semantically equivalent queries are expected to yield consistent recommendations. We used Claude sonnet 5 (Anthropic) to reword the development questions into a held-out test set (Multimedia Appendix 3); patient partners and our team then verified that the rewordings were plausible and consistent. We evaluated recommendation performance (the ability to suggest correct resources) using standard metrics including Precision@K (the proportion of the top K suggestions that were correct), Hit@K (whether at least one was correct), and nDCG@K (whether correct suggestions appeared near the top), for K = 1 to 5 (Multimedia Appendix 4). When a query had more than five ground-truth resources, the additionals were treated as acceptable alternatives, since one query may admit multiple valid recommendations. We assessed across three retrieval methods: sparse (keyword matching), dense (semantic similarity), and hybrid (combining both).

## Results

On the development set, retrieval using keyword matching (sparse) led to the lowest performance (P@1 of 25.0 and a Hit@5 of 43.0 (Table 1)). Using semantic similarity (dense retrieval) increased P@1 to 58.0 and Hit@5 to 86.0. Using both (hybrid retrieval) further improved performance, with a 10.3% increase in P@1 and a 4.7% increase in Hit@5. The best-performing configuration assigned 90% weight to dense retrieval and 10% weight to sparse retrieval, achieving highest performance across nearly all metrics (64.0 P@1, 90.0 Hit@5, and 51.0 nDCG@5).

**Table 1.** Performance across sparse, dense, and hybrid retrieval; *wv* is weight of dense in hybrid retrieval. Test-set results use the best-performing configuration.

| Split | Method | P@1 | P@5 | Hit@5 | nDCG@5 |
| --- | --- | --- | --- | --- | --- |
| Development Set |  |  |  |  |  |
|  | Sparse | 25.0 | 12.2 | 43.0 | 15.2 |
|  | Dense | 58.0 | 38.0 | 86.0 | 44.0 |
| | Hybrid ($wv=0.5$) | 60.0 | 35.4 | 85.0 | 42.3 |
| | Hybrid ($wv=0.6$) | 60.0 | 38.0 | 86.0 | 44.8 |
| | Hybrid ($wv=0.7$) | 59.0 | 37.8 | 87.0 | 44.6 |
| | Hybrid ($wv=0.8$) | 59.0 | 38.6 | 88.0 | 45.1 |
| | Hybrid ($wv=0.9$) | **64.0** | **45.4** | **90.0** | **51.0** |
| Test Set |  |  |  |  |  |
|  | **Hybrid ($wv=0.9$)** | **61.0** | **39.6** | **88.0** | **46.1** |

We then evaluated this best-performing hybrid configuration on the held-out metamorphic test set, to assess if the system could retrieve resources for novel, unseen queries with similar intent. Performance between development and test was close, with a modest decrease of 3-point in P@1 and 2-point in Hit@5. Repeating the test-set evaluation three times produced identical results (Multimedia Appendix 5)

## Discussion

We evaluated a single-pass RAG cancer navigation assistant to find the best way to retrieve resources that answer a user’s question. A hybrid approach, matching keywords but relying mainly on semantic meaning, performed best and generalized well to the held-out test set, giving reproducible recommendations that were robust to rewording. However, while the system reliably included a relevant resource among its top five suggestions (Hit@5 86.0), it was less reliable at ranking the single best resource first (P@1 61.0). For a patient-facing assistant, where users tend to act on the first result they see, this may fall short of clinical needs, though the required accuracy has yet to be established empirically.

Top-rank precision likely lagged because relevance depends on several conditions at once: the patient’s intent, cancer type, service category, and phase of the cancer journey. Semantic similarity can surface resources that meet some but not all of these, leaving several candidates competing for first place. Single-pass RAG also cannot recognize when a question is ambiguous, nor retry when retrieval has failed. Future work will address these limits by adding metadata filtering (e.g., cancer type) and a multi-agent architecture, building on these results to bring us closer to AI-powered navigation ready for clinical use.

## Data Availability

Data access requests can be discussed but are subject to patient privacy considerations

## Data Availability

Available upon request

## Authors’ Contribution

Software: **EH, YZ**

Writing-Original Draft: **EH, YZ, OC, AL, MS, LT, JJN**

Writing-Review & Editing: **All**

