## Supplementary Appendices for "Limits of Single-Pass Retrieval-Augmented Generation for AI-Powered Cancer Care Navigation: A Comparison of Retrieval Strategies": Appendix1.pdf

### Multimedia Appendix 1. Example Test Questions

| Cancer Type | Count | Example |
|---|---|---|
| All | 26 | “My partner had cancer and even though he's better now, he's still dealing with a lot of emotional pain from the experience. Does BC Cancer offer counselling or mental health support for survivors? How do we get him connected to something like that?” |
| Blood and Lymphoid | 6 | "My mom was just told she has terminal leukemia and may only have weeks left. Our whole family is devastated and we want to find out what palliative care options are available to help her." |
| Bone and Soft Tissue | 2 | "I'm meeting with my oncologist next week and I want to be prepared. What treatment options are there for chondrosarcoma?" |
| Breast | 14 | "I had breast cancer surgery last May and ever since I've been dealing with pain and stiffness in my shoulder and arm on that side. It's getting worse and I'm looking for physiotherapy or rehab programs that could help." |
| Central Nervous System | 7 | "I was just diagnosed with a brain tumour — they called it a grade 2 astrocytoma. I've heard about immunotherapy and I'm wondering if it can be used to treat brain cancer." |
| Gastrointestinal | 19 | "I might have esophageal cancer but I haven't been fully diagnosed yet. Where can I find nutritional information and advice for this kind of cancer?" |
| Genitourinary | 12 | "I'm really struggling to accept that I have advanced prostate cancer. I could really use some in-person support — not just something online." |
| Gynecologic | 6 | "I'm scheduled for radiation in my pelvic area and I have so many questions, but my doctor isn't available until August. Is there someone at your organization I could talk to in the meantime?" |
| Head and Neck | 6 | "I just got the results of my salivary gland biopsy and was told I have adenoid cystic carcinoma. I have a couple of questions: Will I need imaging scans to confirm this diagnosis? And what kinds of treatments are available for this?" |
| Lung | 2 | "I'm 59, I smoke, and as far as I know my lungs are fine, but I'm worried about lung cancer. What kind of screening is available for someone in my situation?" |
| Skin | 3 | "I noticed a dark spot on my back recently. Should I be worried that it could be melanoma?" |
