## Supplementary Appendices for "Limits of Single-Pass Retrieval-Augmented Generation for AI-Powered Cancer Care Navigation: A Comparison of Retrieval Strategies": Appendix2.pdf

### Multimedia Appendix 2. Reciprocal Rank Fusion (RRF)

$$s = \frac{w_v}{k + r_v + 1} + \frac{w_b}{k + r_b + 1} \tag{A1}$$

where $r_v$ and $r_b$ denote a document's rank in the vector and BM25 result lists, respectively; $k$ is a smoothing constant; and $w_v$ and $w_b$ are configurable branch weights which we tune to optimize the performance.
