## Supplementary Appendices for "Limits of Single-Pass Retrieval-Augmented Generation for AI-Powered Cancer Care Navigation: A Comparison of Retrieval Strategies": Appendix3.pdf

### Multimedia Appendix 3. Held-out Test Set Generation.

**Prompt:**

"""

This is a set of user queries from cancer patients or caregivers.

Please rephrase or reword them to create a new set of queries while preserving the original meaning. Follow the instructions below while rephrasing the queries:

1. Use the tone and style of a cancer patient or caregiver, ensuring that it doesn't sound like

a medical professional or clinical domain expert.

2. Do not change the original meaning of the query.

3. Do not add any new information to the query that changes the user intent.

4. Do not remove any information from the query that is relevant to the user intent.

"""
