## Supplementary Appendices for "Limits of Single-Pass Retrieval-Augmented Generation for AI-Powered Cancer Care Navigation: A Comparison of Retrieval Strategies": Appendix4.pdf

### Multimedia Appendix 4. Evaluation Metrics

**Precision@K ( p@k)** : For a set of K recommended resources, Precision@K is defined as:

$$\text{Precision@K} = \frac{1}{\min(|G_u|, K)} \sum_{i=1}^{K} \text{rel}_u(i) \tag{1}$$

where $G_u$ is the set of ground-truth relevant resources for query $u$, $|G_u|$ is its cardinality, and $\text{rel}_u(i) \in \{0, 1\}$ is the relevance indicator of the item at rank $i$:

- 1 if the item is relevant;
- 0 otherwise.

**Hit@K** is defined as:

$$\text{Hit@K} = \begin{cases} 1 & \text{if } \exists i \in \{1, \dots, K\} : \text{rel}_u(i) = 1 \\ 0 & \text{otherwise} \end{cases} \tag{2}$$

where $\text{rel}_u(i) \in \{0, 1\}$ is the relevance indicator at rank $i$ as defined above.

**nDCG@K** is defined as:

$$\text{nDCG@K} = \frac{\text{DCG@K}}{\text{IDCG@K}} \tag{3}$$

where the Discounted Cumulative Gain is:

$$\text{DCG@K} = \sum_{i=1}^{K} \frac{\text{rel}_u(i)}{\log_2 (i+1)} \tag{4}$$

and the Ideal DCG is:

$$\text{IDCG@K} = \sum_{i=1}^{\min(|G_u|, K)} \frac{1}{\log_2 (i+1)} \tag{5}$$

with $\text{rel}_u(i) \in \{0, 1\}$ as defined above, and $|G_u|$ the number of ground-truth relevant resources for query $u$.

Higher scores for all metrics indicate better recommendation performance.
