## Supplementary Appendices for "Limits of Single-Pass Retrieval-Augmented Generation for AI-Powered Cancer Care Navigation: A Comparison of Retrieval Strategies": Appendix5.pdf

### Multimedia Appendix 5: Full results

| Split | Method | P@1 | P@2 | P@3 | P@4 | P@5 | Hit@2 | Hit@3 | Hit@4 | Hit@5 | nDCG@2 | nDCG@3 | nDCG@4 | nDCG@5 |
|---|---|---|---|---|---|---|---|---|---|---|---|---|---|---|
| Dev | Sparse retrieval | 25.0 | 20.0 | 15.3 | 13.5 | 12.2 | 31.0 | 33.0 | 39.0 | 43.0 | 21.1 | 17.6 | 16.1 | 15.2 |
| | Dense retrieval | 58.0 | 53.0 | 44.3 | 40.8 | 38.0 | 74.0 | 78.0 | 82.0 | 86.0 | 54.1 | 47.9 | 45.5 | 44.0 |
| | Hybrid (wv=0.5) | 60.0 | 50.5 | 44.7 | 39.5 | 35.4 | 71.0 | 79.0 | 83.0 | 85.0 | 52.6 | 48.2 | 44.8 | 42.3 |
| | Hybrid (wv=0.6) | 60.0 | 54.0 | 47.3 | 42.2 | 38.0 | 74.0 | 81.0 | 83.0 | 86.0 | 55.4 | 50.5 | 47.3 | 44.8 |
| | Hybrid (wv=0.7) | 59.0 | 55.0 | 47.3 | 41.0 | 37.8 | 75.0 | 81.0 | 83.0 | 87.0 | 55.9 | 50.4 | 46.4 | 44.6 |
| | Hybrid (wv=0.8) | 59.0 | 54.0 | 47.0 | 42.0 | 38.6 | 72.0 | 79.0 | 84.0 | 88.0 | 55.1 | 50.1 | 47.0 | 45.1 |
| | Hybrid (wv=0.9) | **64.0** | **56.5** | **51.3** | **47.8** | **45.4** | **74.0** | **81.0** | **86.0** | **90.0** | **58.2** | **54.3** | **52.1** | **51.0** |
| Test | Hybrid (wv=0.9) | 61.0 | 51.5 | 46.3 | 42.2 | 39.6 | 72.0 | 78.0 | 83.0 | 88.0 | 53.6 | 49.6 | 47.1 | 46.1 |
| | Hybrid (wv=0.9) | 61.0 | 51.5 | 46.3 | 42.2 | 39.6 | 72.0 | 78.0 | 83.0 | 88.0 | 53.6 | 49.6 | 47.1 | 46.1 |
| | Hybrid (wv=0.9) | 61.0 | 51.5 | 46.3 | 42.2 | 39.6 | 72.0 | 78.0 | 83.0 | 88.0 | 53.6 | 49.6 | 47.1 | 46.1 |
| | Hybrid (Average) | 61.0 | 51.5 | 46.3 | 42.2 | 39.6 | 72.0 | 78.0 | 83.0 | 88.0 | 53.6 | 49.6 | 47.1 | 46.1 |

**Hit@1 = nDCG@1*
